# Machine Learning Classifier Models for Predicting Sarcopenia in the Elderly Based on Physical Factors

**DOI:** 10.1101/2023.05.03.23288546

**Authors:** Jun-hee Kim

## Abstract

**Background:** As the elderly population gradually increases, musculoskeletal disorders such as sarcopenia are increasing. Diagnostic techniques such as X-ray, CT, and MRI imaging are used to predict and diagnose sarcopenia, and methods using machine learning are gradually increasing.

**Purpose:** The purpose of this study was to create a model that can predict sarcopenia using physical characteristics and activity-related variables without medical diagnostic equipment such as imaging equipment for the elderly aged 60 years or older.

**Method:** A sarcopenia prediction model was constructed using public data obtained from the Korea National Health and Nutrition Examination Survey. Models were built using the multi-layer perceptron, XGBoost, LightGBM, and RandomForest algorithms, and the feature importance of the model with the highest accuracy was analyzed through evaluation metrics.

**Result:** The sarcopenia prediction model built with the LightGBM algorithm showed the highest test accuracy at 0.852. In constructing the LightGBM model, physical characteristics variables such as BMI showed high importance, and activity-related variables were also used in constructing the model.

**Conclusion:** The sarcopenia prediction model composed only of physical characteristics and activity-related factors showed excellent performance, and the use of this model will help predict sarcopenia in the elderly living in communities with insufficient medical resources or difficult access to medical facilities.

## I. INTRODUCTION

A decrease in skeletal muscle and consequent decrease in muscle strength can be seen as a representative physical change in the elderly.(1) The amount of skeletal muscle decreases by about 8% every 10 years (annual average of 0.8%) from the age of 40, and after the age of 70, more rapid muscle loss occurs, and it appears to decrease by about 15% for 10 years.(1) As such, rapid changes in body composition and body functions, such as muscle loss due to aging, can be defined as sarcopenia.(1) The prevalence of sarcopenia shows a tendency to gradually increase with increasing age.(2) In Korea, 13.0% of males and 21.7% of females in the middle-aged (40 to 59 years) had sarcopenia, and 21.6% of males and 30.7% of females in the elderly (60 or older) had sarcopenia.(2)

The clinical significance of sarcopenia can be summarized as a decrease in muscle strength due to a decrease in muscle mass, and an accompanying increase in physical disability and mortality. Sarcopenia has been reported to be closely related to physical dysfunction, and in particular, problems such as decreased gait function or increased risk of falling have been reported.(3,4) In addition, sarcopenia was often accompanied by diseases such as metabolic dysfunction or geriatric chronic diseases, and was also considered to be the cause of these diseases.(5,6) Therefore, sarcopenia due to aging directly causes muscle strength deterioration, leading to deterioration of physical function, and may cause or accompany chronic disabilities, and may increase the risk of death in the elderly.(7)

According to the definition of sarcopenia, measurement of absolute muscle mass is the best assessment indicator for diagnosing sarcopenia.(8) Therefore, examination using imaging measures such as MRI or CT to measure absolute muscle mass is currently considered the gold standard for diagnosing sarcopenia.(9) These two imaging techniques show the highest accuracy and reproducibility, and based on high image resolution, it is possible to selectively measure only muscles excluding fat among tissues composing the body.(10) However, imaging using MRI or CT is expensive and requires skilled operator experience and skill.(10) In addition, patients require high compliance with MRI or CT imaging, and in the case of CT, patients may be exposed to high levels of radiation.(10) For this reason, in clinical practice, sarcopenia is diagnosed using dual-energy X-ray absorptiometry (DXA), which has a low radiation exposure and is relatively easy to measure.(9,10) DXA can measure body composition by classifying it into bone mineral content, body fat mass, and lean body mass, and can measure it separately for each limb and trunk.(10) The sum of the muscle mass excluding the bone mineral mass from the lean body mass of both limbs is called the appendicular skeletal muscle mass (ASM) and is used as a major indicator in the diagnosis of sarcopenia.(11) If even these diagnostic methods are difficult to apply, sarcopenia can be diagnosed with bioelectrical impedance analysis (BIA).(12) In addition to diagnosing sarcopenia using these devices, attempts have been made to prevent or improve sarcopenia by more simply and easily diagnosing sarcopenia through functional tests such as grip strength and walking speed.(13,14) However, studies diagnosing sarcopenia using simple functional tests, such as grip strength, appear differently depending on the race of subjects, or the methods of measuring functional factors used in studies have not yet been standardized or consistent.(15,16)

Recently, to overcome the limitations of assessments for diagnosing sarcopenia, methods for diagnosis through models using machine learning have been studied and presented. Machine learning is a technique that can predict a specific value or classification by using a large amount of data containing various variables.(17) A machine learning model that diagnoses image data through clinical measurement equipment instead of an expert is being built to improve the accuracy of diagnosing sarcopenia, and it is also possible to predict sarcopenia using imaging data from abdomen or chest rather than limbs.(18–20) In addition, it is possible to diagnose sarcopenia or predict the risk by modeling variables that can be related, such as physical characteristics, nutrition, and blood-related variables, using machine learning, rather than analyzing only a few variables such as muscle strength or walking speed.(21,22) These studies suggested that the prediction of sarcopenia using a machine learning model would be more efficient in terms of time and cost than the traditional method of diagnosing sarcopenia using imaging measurements.(19,21)

Although these predictors are said to be efficient in terms of time and cost, to measure these variables, medical devices or precise examinations by experts are required. This suggests that it may be difficult to perform assessments according to the elderly living in underdeveloped facilities with poor access to medical facilities or the rapidly growing elderly population.(8) Therefore, a model that can be used as a screening or predict the risk of sarcopenia before using medical equipment or performing a detailed examination by an expert to diagnose sarcopenia in the elderly will be needed. Such a model should be composed of variables that are related to sarcopenia and are easy to measure, such as physical characteristics and activities.(23,24)

Therefore, the purpose of this study was to create a predictive model for diagnosing sarcopenia based on easily measurable variables such as physical characteristics and activity-related variables. To build this predictive model, the following steps were required. First, it was to find out which variables are needed to build models among variables related to physical characteristics and activities. Second, it was to build several models with selected variables and improve performance through hyperparameter tuning. Third, it was to compare various models and select the model with the best performance among them. Fourth, it was to find out which variables had the highest importance in constructing the best performing model.

## II. METHODS

### 1. Data Source

The flow of this research process is shown in Figure 1. Data from the Korea National Health and Nutrition Examination Survey (KNHANES) conducted by the Korea Disease Control and Prevention Agency from 2008 to 2011 were used in this study. A total of 37,753 samples were investigated during that period. Among the data samples, there were a total of 5,573 data samples with dual energy x-ray data, age 60 years or older, and no missing responses to physical characteristics and activity-related variables. Data samples used in the study were provided with permission from the Korea Disease Control and Prevention Agency as data with personal identification information deleted.

**Figure 1.**
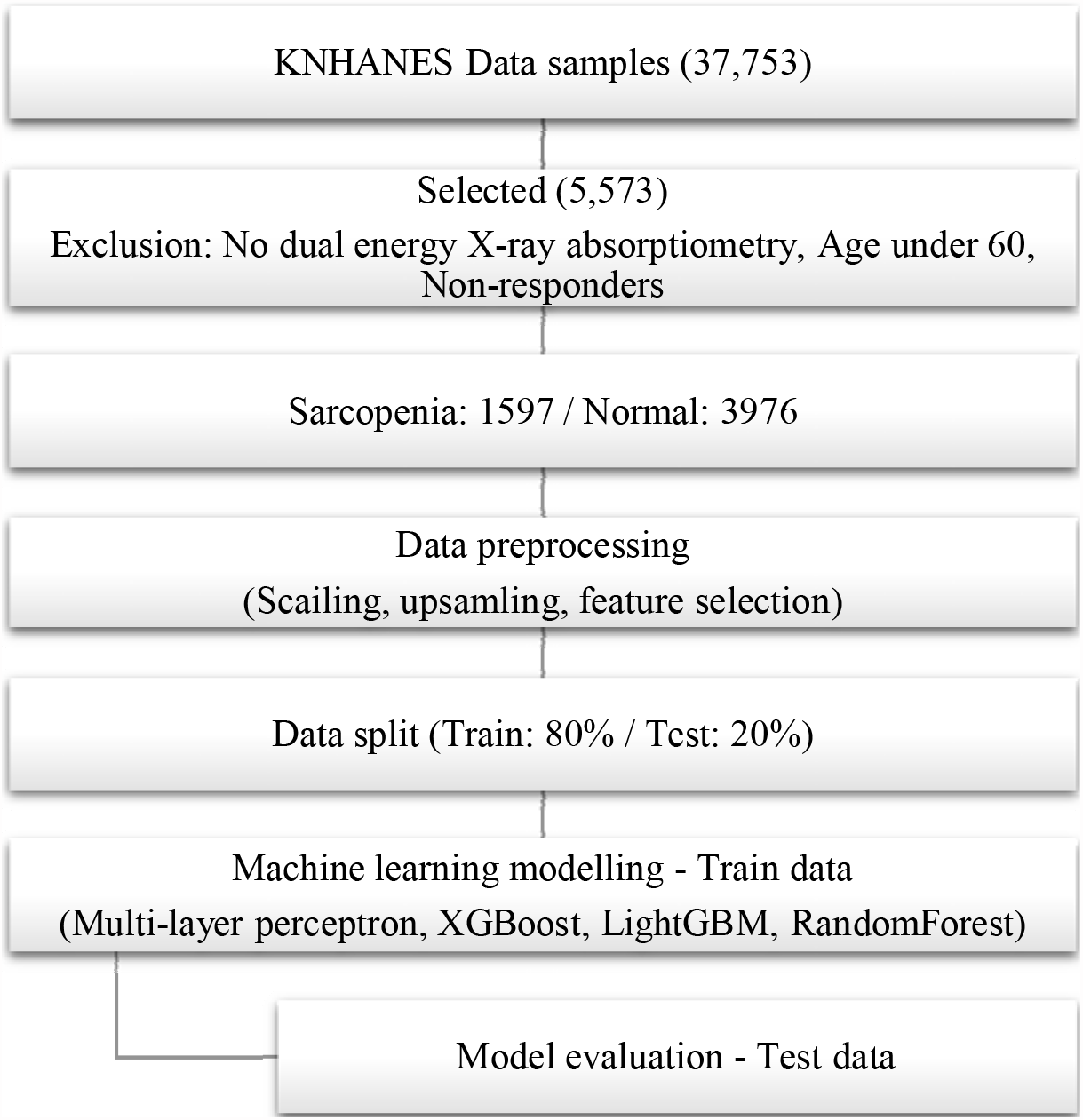
Flowchart of the study.

### 2. Data Preprocessing

This study used variables reflecting physical characteristics such as age, sex, height, weight and BMI, and variables reflecting the degree of physical activity such as working hours or exercise participation time per week. Table 1 shows the variables and descriptions of the data samples used in this study. Categorical variables were quantified through LabelEncoder, and numerical variables were standardized using StandardScaler. To avoid overfitting and poor performance due to imbalanced data, SMOTE was used to randomly generate data samples to match the proportions between the data. LASSO algorithm was used to select features to use the multi-layer perceptron (MLP) model. And RFECV (Recursive Feature Elimination with Cross-Validation) was used for feature selection of XGBoost, LightGBM, and RandomForest models. Finally, before training the models, the entire dataset was divided into a training dataset and a test dataset at a ratio of 8:2.

**Table 1.** Description of variables.

| **KNHANES** | **Description** | **Sarcopenia** | **Normal** |
| --- | --- | --- | --- |
| age | Age | 70.76 ± 6.17 | 68.54 ± 5.84 |
| sex | Gender | M:941, F:656 | M:1467, F:2509 |
| HE_ht | Height | 159.03 ± 8.85 | 157.03 ± 8.89 |
| HE_wt | Weight | 54.09 ± 8.31 | 61.39 ± 9.95 |
| HE_wc | Waist circumference | 78.88 ± 8.61 | 86.02 ± 8.87 |
| BMI | Body mass index | 21.34 ± 2.41 | 24.81 ± 2.94 |
| BO1_1 | Weight change | N:1170, I:353, D:74 | N:2980, I:676, D:320 |
| EQ5D | Quality of life | 0.86 ± 0.18 | 0.87 ± 0.17 |
| EC_wht_23 | Average working hours per week | 16.45 ± 24.77 | 18.53 ± 25.5 |
| BE3_11 | Vigorous physical activity per week | 1.58 ± 1.63 | 1.76 ± 1.78 |
| BE3_21 | Moderate physical activity per week | 2.12 ± 2.17 | 2.56 ± 2.45 |
| BE3_31 | Walking days per week | 5.33 ± 2.82 | 5.27 ± 2.79 |
| LQ4_00 | Activity restriction | Y:537, N:1060 | Y:1217, N:2759 |
| EC1_1 | Occupational activity | Y:605, N:992 | Y:1704, N:2272 |

\*M, male; F, female; Y, yes; N, no; I, increase; D, decrease

### 3. Machine learning algorithms

A sarcopenia prediction model was developed using four machine learning algorithms: MLP, XGBoost, LightGBM, and RandomForest. This study used a MLP model with one input layer, one output layer and five hidden layers with varying numbers of nodes. The ReLU activation function was used for the hidden layers, while the sigmoid activation function was used for the output layer. Dropout ratio and batch normalization were applied to prevent overfitting, and the Adam optimizer was used for gradient descent. The data was split with a 0.2 validation ratio, and EarlyStopping was used to terminate learning when the validation loss did not improve for 20 epochs. The other three models used the StratifiedKFold for cross-validation to ensure unbiased performance. This technique partitions the data into K folds and ensures that each fold contains approximately equal proportions of samples from each class. This allows the model to be trained and evaluated on a representative subset of the data to obtain a more accurate performance estimate. In this study, a 5-fold training data set was created for each model, and the model was built by repeating training and validation five times. The hyperparameters of each model were then tuned using the GridSearchCV technique.

### 4. Model evaluation

The model’s performance was assessed by comparing its predicted and actual classes using a confusion matrix that includes TP (true positive), FP (false positive), FN (false negative), and TN (true negative) values, based on test data. The accuracy, precision, recall, and F1-score were calculated from the matrix to evaluate the models. In addition, models were compared using the ROC AUC curve. Through the feature importance of the model with the highest accuracy, important factors in generating the model were analyzed.

## III. RESULTS

### 1. Feature selection

All the mean test scores of the models according to variable selection using RFECV and LASSO exceeded 0.75. The variables selected and excluded for each algorithm to build the predictive models for sarcopenia diagnosis, and the mean test scores are shown in Table 2.

**Table 2.**
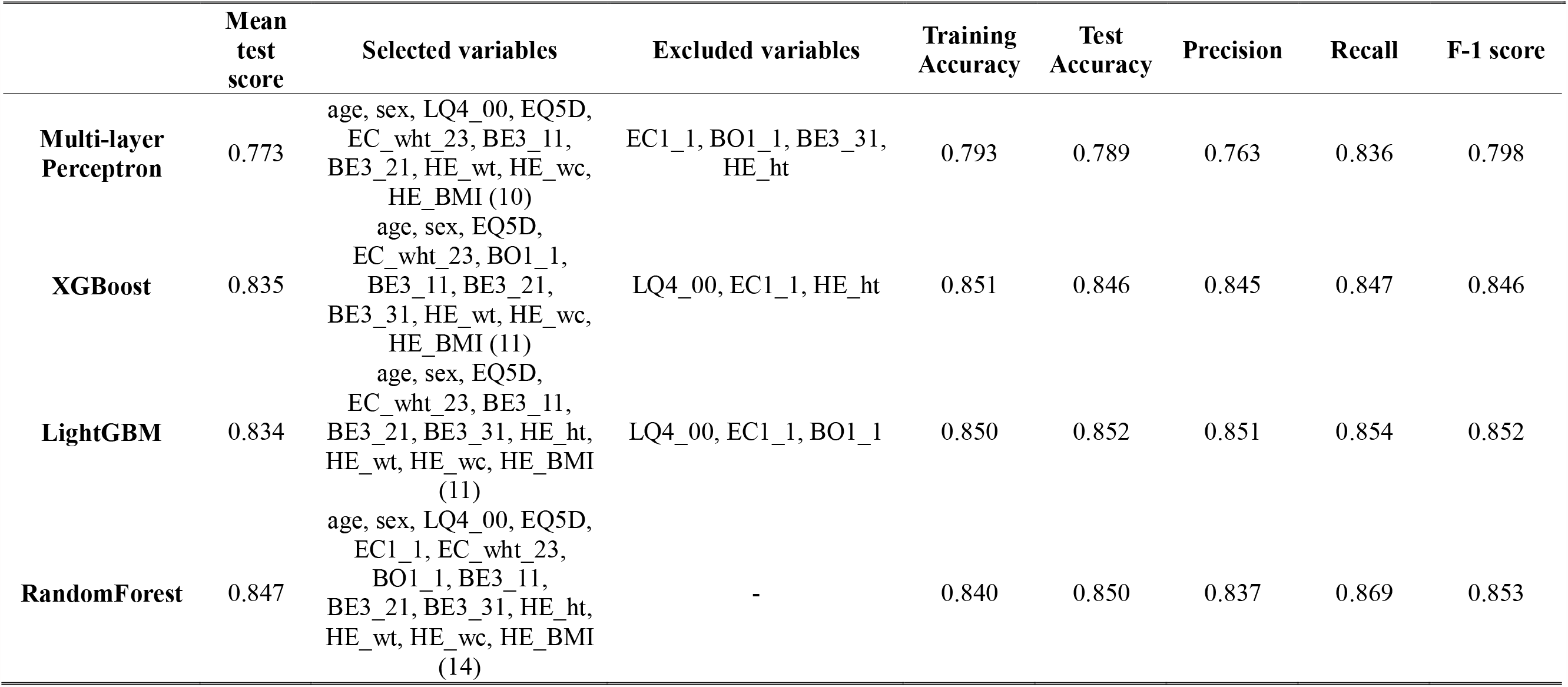
Feature selection and model evaluation scores.

### 2. Model performance

Figure 2 shows the confusion matrix created with test data to evaluate the performance of models built with MLP, XGBoost, LightGBM, and RandomForest. Based on confusion matrix, the MLP model showed the lowest performance with an accuracy of 0.79 when evaluated with test data. The model with the highest performance was the model composed of the LightGBM algorithm. The test accuracy of this model was 0.852, showing slightly higher performance than models using XGBoost and RandomForest algorithms. Table 2 shows the accuracy, precision, recall, and F1-score of the models using the multi-layer perceptron, XGBoost, LightGBM, and RandomForest algorithms. The models compared through the ROC-AUC curve showed similar high performance except for the MLP model (Figure 2).

**Figure 2.**
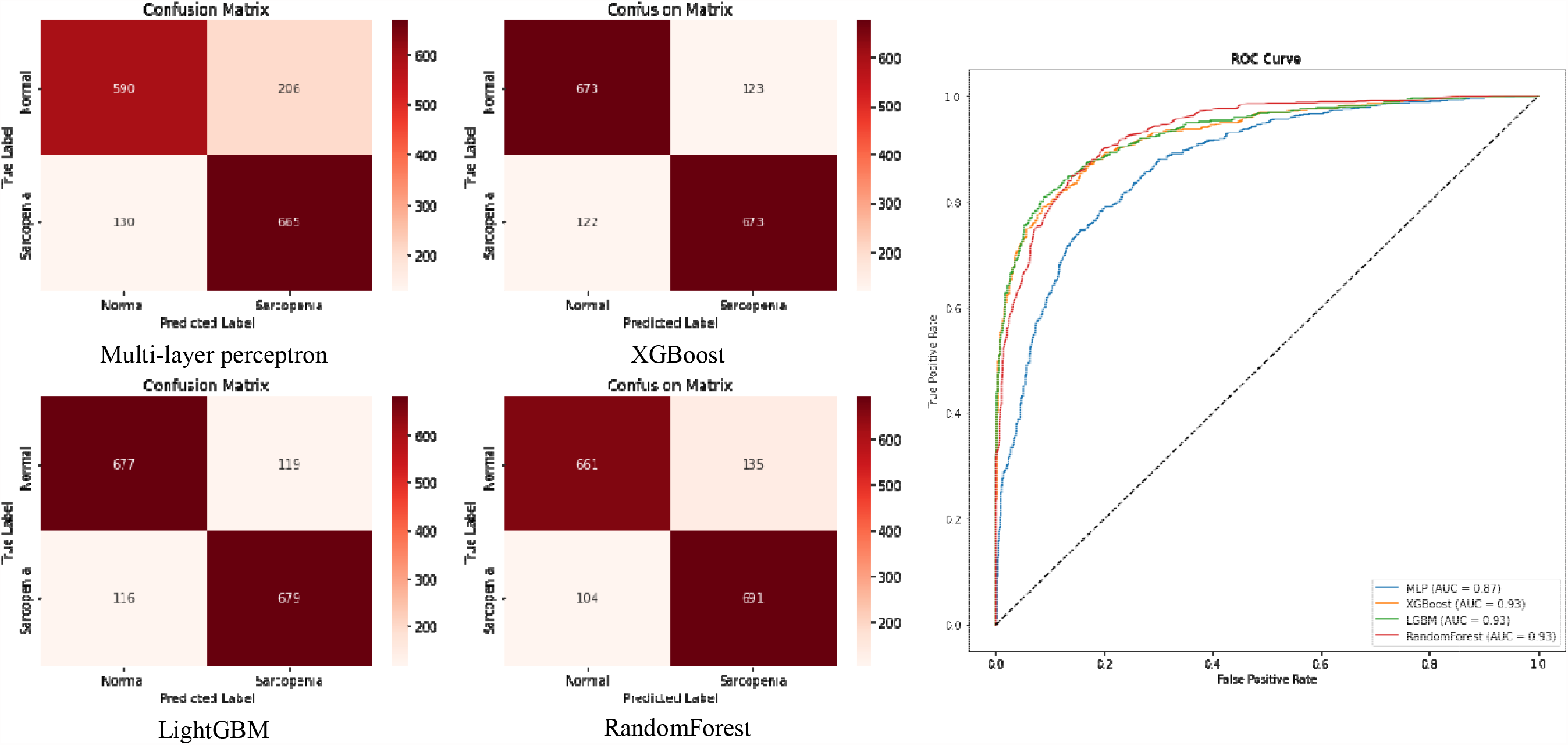
Confusion matrix and ROC-AUC curve of sarcopenia prediction model.

### 3. Feature importance

The variable with the highest importance in constructing the LightGBM model in predicting sarcopenia was BMI Figure 3). In addition, variables related to individual physical characteristics such as age, waist circumference, height, and weight were found to be important variables in constructing a sarcopenia prediction model. EQ-5D, which represents health-related quality of life, was found to be an important variable in model construction next to variables representing individual physical characteristics, and variables such as the number of hours of work participation per week were the next most important variables, followed by the number of days of participation in walking or moderate- and high-intensity exercise. Gender appeared to be the least important variable in constructing the sarcopenia prediction model.

**Figure 4.**
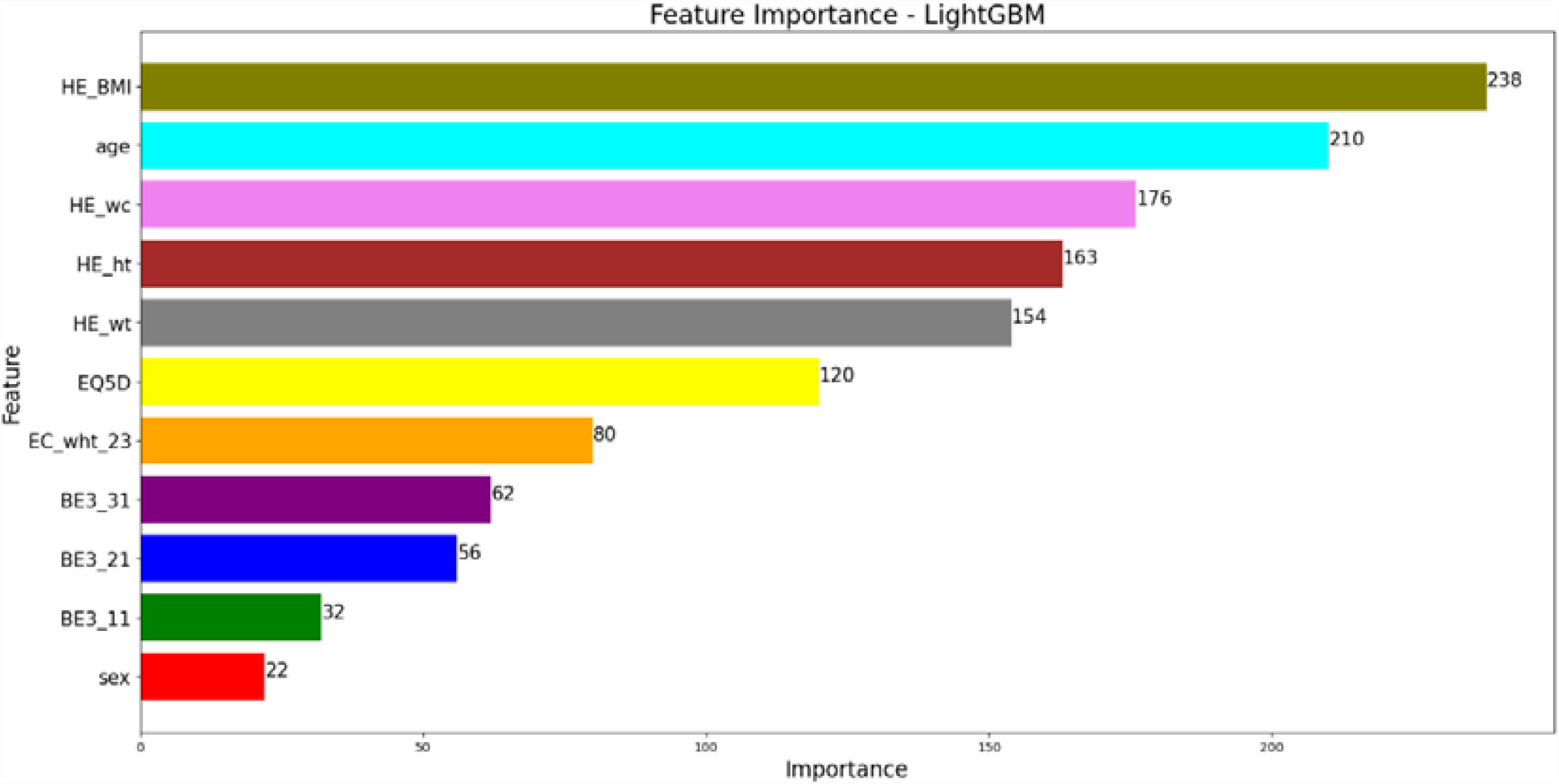
Feature importance of LightGBM model.

## IV. DISCUSSION

The purpose of this study was to create a model to predict the diagnosis of sarcopenia in the elderly with relatively easy-to-obtain physical characteristics and activity-related variables compared to variables measured by experts with medical equipment such as DXA to measure lean muscle mass. The sarcopenia prediction model constructed in this study showed an accuracy performance of about 85% or more, except for the MLP model. In constructing a high-performance model, variables related to individual physical characteristics such as BMI and age were the most important, followed by variables related to exercise participation or occupational activity.

The machine learning model built with the data measured by the diagnostic imaging equipment showed a relatively high level of accuracy. Burns al. (2020) built a machine learning model to predict sarcopenia using abdominal cross-sectional CT taken at the lumbar level of an elderly person, and the accuracy (expressed as AUC score) of this model was around 94%.(19) Ryu et al. (2022) built an model to predict the prevalence of sarcopenia using chest X-ray data in a simpler way than CT scan, and the accuracy (expressed as AUC score)of this model was 74-88%.(20) In addition, even when the machine learning model was constructed with variables other than those measured by imaging equipment, it showed high performance in diagnosing sarcopenia. Ko et al. (2021) developed a model with 81-88% accuracy by analyzing patterns during timed-up-and-go (TUG) and 6-minute walk test (6mWT) by machine learning using an IMU sensor.(25) Kang et al. (2019) created a model to predict sarcopenia with 78-82% accuracy (expressed as AUC score) using physical variables such as BMI and age, blood-related variables such as red blood cell count and white blood cell count, and dietary variables such as fiber intake, fat intake, and protein intake.(21) Similarly, the AUC score of the sarcopenia prediction model using body characteristics and physical activity variables in this study was 87-93%. In addition, the accuracy reached about 85% for three of the four models. The performance of the sarcopenia prediction model generated through this study did not show outstanding performance compared to other models. Nevertheless, there are several advantages compared to sarcopenia predictive models in other studies. First, the variables used to build the model in this study were factors that did not require imaging techniques using expensive medical equipment such as X-ray, CT, and MRI. Second, the variables in this study did not require invasive testing for human specimens, such as blood-related factors or muscle biopsies. Third, the variables used in model construction were not variables that required high-level skills to measure or that had to be measured by experts. Considering these advantages, even if the examiners or investigators do not have high-level skills, they will be able to measure the data used in this model, and even the subjects will be able to measure themselves using scales or questionnaires. In addition, the subject can be remotely It may be possible to input data and remotely receive sarcopenia prediction results for these data.

Variables related to physical characteristics showed high importance in constructing the LGBM model, which showed the highest accuracy among the models in this study. BMI showed the highest feature importance, followed by age, waist circumference, height, and weight. In the case of sarcopenia in Asian elderly with diabetes, it was found that sarcopenia decreased significantly as BMI increased.(26) In addition, according to the model for predicting sarcopenia constructed with the RandomForest algorithm by Kim et al (2019), BMI served as the first variable to classify the elderly over 60 as sarcopenia and normal.(27) According to the deep learning model built by Somasundaram et al (2022) for the diagnosis of sarcopenia in adolescents, the skeletal muscle area measured by abdominal CT at the L3 level, one of the markers of sarcopenia diagnosis, showed a high correlation (0.75-0.94) with weight, height, age, and BMI in adolescence.(28) Based on these studies, the model constructed in our study would also have shown high importance in constructing the model for physical characteristics variables. In addition, studies related to sarcopenia and quality of life have been conducted. According to the review article, 4 studies measuring quality of life with the Short-form General Health Survey (SF-36) and 2 studies using the EQ-5D instrument, Subjects with sarcopenia were found to have significantly higher rates of problems related to quality of life.(29) For this reason, in the model of this study, EQ-5D would have shown the highest importance next to physical characteristics. The feature importance of physical activity-related variables in the model created in this study was lower than that of physical characteristics or quality of life variables. Nevertheless, the importance of physical activity in preventing muscle loss has been emphasized.(30) There is also evidence that physical activity, a modifiable lifestyle behavior, can partially reverse age-related skeletal muscle dysfunction.(30,31) Therefore, variables related to physical activity can be used as predictive variables in constructing a sarcopenia prediction model even if the feature importance is not high.

There are several limitations in this study. First, the sarcopenia predictive model of this study was constructed using data of the elderly over 60 years of age, so the performance of this model cannot be guaranteed for predicting sarcopenia in adolescents or young people. Second, in constructing the predictive model in this study, the model may have been overfitted because upsampling was used to balance the number of sarcopenia and normal data. Third, according to the inherent characteristics of machine learning models, it was possible to infer the correlation of variables related to sarcopenia prediction by calculating the feature importance in model creation, but it was difficult to interpret it as a causal relationship. Future studies will need to build a model by additionally collecting data from various age groups, and additionally discover several variables that are expected to have a high impact on sarcopenia and use them to construct a model. In addition, research to identify the causal relationship between variables representing high feature importance and sarcopenia through a machine learning model should be conducted.

## V. CONCLUSION

In this study, a sarcopenia prediction model was created using physical characteristics and activity variables of sarcopenia and normal elderly over 60 years of age. The performance of the model showed an accuracy of about 85%, and physical characteristics variables such as BMI, age, height, and weight were found to be important in constructing this model. In addition, through feature importance, it was found that activity-related variables such as EQ-5D, occupational activity, and number of days participating in physical activity were also used to construct the model. Since this model was constructed using relatively easy-to-measure variables without using medical equipment, it will be able to help predict and prevent sarcopenia in the elderly in areas with insufficient medical resources or poor access to medical facilities.

## Data Availability

All data produced are available online at Korea National Health and Nutrition Examination Survey (KNHANES)

https://knhanes.kdca.go.kr/knhanes/sub03/sub03_02_05.do

## Conflicts of Interest Statement

The author whose name is listed immediately below certifies that they have NO affiliations with or involvement in any organization or entity with any financial interest (such as honoraria; educational grants; participation in speakers’ bureaus; membership, employment, consultancies, stock ownership, or other equity interest; and expert testimony or patent-licensing arrangements), or non-financial interest (such as personal or professional relationships, affiliations, knowledge, or beliefs) in the subject matter or materials discussed in this manuscript. I understand that the Corresponding Author is the sole contact for the Editorial process and is responsible for submissions of revisions and final approval of proofs.

Author name: Jun-hee Kim

## VI. References

1. Mitchell WK, Williams J, Atherton P, Larvin M, Lund J, Narici M. Sarcopenia, dynapenia, and the impact of advancing age on human skeletal muscle size and strength; a quantitative review. Front Physiol. 2012;3:260.

2. Oh JM, Choi J-H, Lee YJ, Lee YR, Youn NH, Song HJ. Association between sarcopenia and health-related quality of life in Korean adults: based on the Fifth Korean National Health and Nutrition Examination Survey (2010-2011). Korean J Fam Pract. 2017;7(6):870–6.

3. Woo N, Kim SH. Sarcopenia influences fall-related injuries in community-dwelling older adults. Geriatr Nurs (Minneap). 2014;35(4):279–82.

4. PerezlJSousa MA, VenegaslJSanabria LC, ChavarrolJCarvajal DA, CanolJGutierrez CA, Izquierdo M, CorrealJBautista JE, et al. Gait speed as a mediator of the effect of sarcopenia on dependency in activities of daily living. J Cachexia Sarcopenia Muscle. 2019;10(5):1009–15.

5. Biolo G, Cederholm T, Muscaritoli M. Muscle contractile and metabolic dysfunction is a common feature of sarcopenia of aging and chronic diseases: from sarcopenic obesity to cachexia. Clin Nutr. 2014;33(5):737–48.

6. Lim H-S, Park Y-H, Suh K, Yoo MH, Park HK, Kim HJ, et al. Association between sarcopenia, sarcopenic obesity, and chronic disease in Korean elderly. J Bone Metab. 2018;25(3):187.

7. Chang S, Lin P. Systematic literature review and metalJanalysis of the association of sarcopenia with mortality. Worldviews EvidencelJBased Nurs. 2016;13(2):153–62.

8. Beaudart C, Rizzoli R, Bruyère O, Reginster J-Y, Biver E. Sarcopenia: burden and challenges for public health. Arch public Heal. 2014;72(1):1–8.

9. Albano D, Messina C, Vitale J, Sconfienza LM. Imaging of sarcopenia: old evidence and new insights. Eur Radiol. 2020;30:2199–208.

10. Guglielmi G, Ponti F, Agostini M, Amadori M, Battista G, Bazzocchi A. The role of DXA in sarcopenia. Aging Clin Exp Res. 2016;28:1047–60.

11. Belarmino G, Gonzalez MC, Sala P, Torrinhas RS, Andraus W, D’Albuquerque LAC, et al. Diagnosing sarcopenia in male patients with cirrhosis by duallJenergy XlJray absorptiometry estimates of appendicular skeletal muscle mass. J Parenter Enter Nutr. 2018;42(1):24–36.

12. Gonzalez MC, Barbosa-Silva TG, Heymsfield SB. Bioelectrical impedance analysis in the assessment of sarcopenia. Curr Opin Clin Nutr Metab Care. 2018;21(5):366–74.

13. Yuki A, Ando F, Shimokata H. Transdisciplinary approach for sarcopenia. Sarcopenia: definition and the criteria for Asian elderly people. Clin Calcium. 2014;24(10):1441–8.

14. Reijnierse EM, De Van Der Schueren MAE, Trappenburg MC, Doves M, Meskers CGM, Maier AB. Lack of knowledge and availability of diagnostic equipment could hinder the diagnosis of sarcopenia and its management. PLoS One. 2017;12(10):e0185837.

15. Ha Y-C, Hwang S-C, Song S-Y, Lee C, Park K-S, Yoo J-I. Hand grip strength measurement in different epidemiologic studies using various methods for diagnosis of sarcopenia: a systematic review. Eur Geriatr Med. 2018;9:277–88.

16. Cawthon PM, Travison TG, Manini TM, Patel S, Pencina KM, Fielding RA, et al. Establishing the link between lean mass and grip strength cut points with mobility disability and other health outcomes: proceedings of the sarcopenia definition and outcomes consortium conference. Journals Gerontol Ser A. 2020;75(7):1317–23.

17. Jordan MI, Mitchell TM. Machine learning: Trends, perspectives, and prospects. Science. 2015;349(6245):255–60.

18. Kim YJ. Machine learning models for sarcopenia identification based on radiomic features of muscles in computed tomography. Int J Environ Res Public Health. 2021;18(16):8710.

19. Burns JE, Yao J, Chalhoub D, Chen JJ, Summers RM. A machine learning algorithm to estimate sarcopenia on abdominal CT. Acad Radiol. 2020;27(3):311–20.

20. Ryu J, Eom S, Kim HC, Kim CO, Rhee Y, You SC, et al. Chest XlJraylJbased opportunistic screening of sarcopenia using deep learning. J Cachexia Sarcopenia Muscle. 2023;14(1):418–28.

21. Kang Y-J, Yoo J-I, Ha Y. Sarcopenia feature selection and risk prediction using machine learning: A cross-sectional study. Medicine (Baltimore). 2019;98(43).

22. Wu L-W, OuYoung T, Chiu Y-C, Hsieh H-F, Hsiu H. Discrimination between possible sarcopenia and metabolic syndrome using the arterial pulse spectrum and machine-learning analysis. Sci Rep. 2022;12(1):21452.

23. Beaudart C, Dawson A, Shaw SC, Harvey NC, Kanis JA, Binkley N, et al. Nutrition and physical activity in the prevention and treatment of sarcopenia: systematic review. Osteoporos Int. 2017;28:1817–33.

24. Kim KM, Jang HC, Lim S. Differences among skeletal muscle mass indices derived from height-, weight-, and body mass index-adjusted models in assessing sarcopenia. Korean J Intern Med. 2016;31(4):643.

25. Ko JB, Kim KB, Shin YS, Han H, Han SK, Jung DY, et al. Predicting Sarcopenia of Female Elderly from Physical Activity Performance Measurement Using Machine Learning Classifiers. Clin Interv Aging. 2021;1723–33.

26. Fukuoka Y, Narita T, Fujita H, Morii T, Sato T, Sassa MH, et al. Importance of physical evaluation using skeletal muscle mass index and body fat percentage to prevent sarcopenia in elderly Japanese diabetes patients. J Diabetes Investig. 2019;10(2):322–30.

27. Kim S, Yi C, Lim J. Risk Factors for Sarcopenia, Sarcopenic Obesity, and Sarcopenia Without Obesity in Older Adults. Phys Ther Korea. 2021;28(3):177–85.

28. Somasundaram E, Castiglione JA, Brady SL, Trout AT. Defining normal ranges of skeletal muscle area and skeletal muscle index in children on CT using an automated deep learning pipeline: implications for sarcopenia diagnosis. Am J Roentgenol. 2022;219(2):326–36.

29. Tsekoura M, Kastrinis A, Katsoulaki M, Billis E, Gliatis J. Sarcopenia and its impact on quality of life. GeNeDis 2016 Genet Neurodegener. 2017;213–8.

30. Pillard F, Laoudj-Chenivesse D, Carnac G, Mercier J, Rami J, Rivière D, et al. Physical activity and sarcopenia. Clin Geriatr Med. 2011;27(3):449–70.

31. Bosaeus I, Rothenberg E. Nutrition and physical activity for the prevention and treatment of age-related sarcopenia. Proc Nutr Soc. 2016;75(2):174–80.

